# Evaluation of the INDICAID™ COVID-19 Rapid Antigen Test in symptomatic populations and asymptomatic community testing

**DOI:** 10.1101/2021.05.26.21257063

**Authors:** Ricky Y. T. Chiu, Noah Kojima, Garrett Lee Mosley, Kwok Kin Cheng, David Y. Pereira, Matthew Brobeck, Tsun Leung Chan, Jonpaul Sze-Tsing Zee, Harsha Kittur, Tenny Cheuk-Yiu Chung, Eric Tsang, Kajal Maran, Raymond Wai-Hung Yung, Alex Chin-Pang Leung, Ryan Ho-Ping Siu, Jessica Pui-Ling Ng, Tsz Hei Choi, Mei Wai Fung, Wai Sing Chan, Ho Yin Lam, Koon Hung Lee, Sean Parkin, Felix C. Chao, Stephen Ka-Nung Ho, Daniel R. Marshak, Edmond Shiu-Kwan Ma, Jeffrey D. Klausner

**Author notes:** **Correspondence:** N. Kojima, Department of Medicine at UCLA, 10833 Le Conte Ave, Los Angeles, CA 90095.

## Abstract

**Background:** As the COVID-19 pandemic continues to cause substantial morbidity and mortality, there is an increased need for rapid, accessible assays for SARS-CoV-2 detection.

**Methods:** Here we present a clinical evaluation and real-world implementation of the INDICAID™ COVID-19 Rapid Antigen Test (INDICAID™ Rapid Test). A multi-site clinical evaluation of the INDICAID™ Rapid Test using prospectively collected samples from symptomatic subjects was performed. The INDICAID™ Rapid Test was then implemented at COVID-19 outbreak screening centers in Hong Kong to screen individuals for COVID-19 to prioritize confirmatory RT-PCR testing among asymptomatic populations.

**Results:** The clinical evaluation in symptomatic patient populations demonstrated a positive percent agreement and negative percent agreement of 85.3% (95% confidence interval [95% CI]: 75.6% - 91.6%) and 94.9% (95% CI: 91.6% - 96.9%), respectively, when compared to laboratory-based RT-PCR testing. When used during outbreak testing of 22,994 asymptomatic patients, the INDICAID™ Rapid Test demonstrated a positive percent agreement of 84.2% (95% CI: 69.6% - 92.6%) and a negative percent agreement of 99.9% (95% CI: 99.9% - 100%) compared to laboratory-based RT-PCR testing. When incorporated in a testing algorithm, the INDICAID™ Rapid Test reduced time to confirmatory positive result from an average 10.85 hours (standard RT-PCR only) to 0.84 hours, depending on the algorithm.

**Conclusion:** The INDICAID™ Rapid Test has excellent performance when compared to laboratory-based RT-PCR testing, and when used in tandem with RT-PCR, reduces the time to confirmatory positive result.

**Summary:** In clinical evaluations, the INDICAID™ COVID-19 Rapid Antigen Test demonstrated high sensitivity and specificity in symptomatic and asymptomatic patient populations. When used in tandem with RT-PCR testing, the INDICAID™ Rapid Test expedited confirmatory results and may help reduce SARS-CoV-2 outbreaks.

## INTRODUCTION

The global SARS-CoV-2 pandemic has continued despite implementation of significant public health measures [1]. Over 155 million worldwide cases of COVID-19 and over 3 million COVID-19 deaths have been reported as of May 2021. Rapid identification of SARS-CoV-2 infection, patient isolation, and contact tracing are essential for disease containment [2].

The current gold standard for detecting SARS-CoV-2 is reverse-transcriptase polymerase chain reaction (RT-PCR) [3]. While RT-PCR can detect nucleic acids from SARS-CoV-2 with high sensitivity, RT-PCR requires equipment, special training, and can take days until results are available following sample collection [4]. Due to the uniquely high transmissibility of SARS-CoV-2 (basic reproductive number R_0_ of 2.87), long turnaround times for results may lead to a high number of avoidable transmissions [5][6].

In contrast, lateral flow immunoassays (LFAs) are an inexpensive testing solution that can be used at point-of-care settings, do not require laboratory equipment, and can generate results quickly. However, the performance of LFA-based SARS-CoV-2 rapid antigen tests in community testing settings can vary significantly [7–9].

In this study, we evaluated the clinical performance of the LFA-based INDICAID™ COVID-19 Rapid Antigen Test (INDICAID™ Rapid Test) by PHASE Scientific International Ltd. A prospective multi-site clinical study was performed in symptomatic patient populations in point-of-care (POC) community testing sites in the United States. The performance of the INDICAID™ Rapid Test was also evaluated in COVID-19 outbreak screening centers in Hong Kong as a part of an algorithm testing approach (termed “Dual-Track”) to screen for COVID-19 positive patients prior to RT-PCR testing in asymptomatic patient populations.

## METHODS

### Description of the INDICAID™ COVID-19 Rapid Antigen Test

The INDICAID™ Rapid Test by PHASE Scientific is a LFA designed for the qualitative detection of SARS-CoV-2 nucleocapsid protein in nasal swab samples. The test produces a simple readout within 20 minutes with the presence of a visible test line to indicate detection of the SARS-CoV-2 antigen. In contrast to other reader-based rapid antigen platforms, the INDICAID™ Rapid Test achieves results without any additional equipment, power source, or training.

#### Sample collection and procedure of the INDICAID™ COVID-19 Rapid Antigen Test

To conduct an INDICAID™ Rapid Test, a nasal swab sample is collected by inserting the provided swab 1 inch into the nasal cavity. The swab is rubbed against the inside walls of both nostrils 5 times in a large circular path. The swab is then dipped into a buffer solution to elute the sample. Finally, three drops of the buffer solution-specimen mix are applied to the LFA test device. After 20 minutes, the user observes the test device for the presence or absence of a test line that indicates detection of the SARS-CoV-2 antigen. An Internal Quality Control line is included to indicate whether the test has been performed correctly.

### Prospective multi-site clinical evaluation of INDICAID™ COVID-19 Rapid Antigen Test

#### Populations and study locations

Between November 30, 2020 and January 8, 2021, study participants were enrolled at two US clinical sites, CityHealth Urgent Care San Francisco, CA and CityHealth Urgent Care Oakland, CA.

As part of the inclusion criteria, study participants were required to be at least 5 years of age and report onset of at least two of the following COVID-19 symptoms within 5 days or less: fever or chills, fatigue, sore throat, congestion or runny nose, cough, headache, diarrhea, shortness of breath or difficulty breathing, muscle or body aches, new loss of taste or smell, nausea, or vomiting. A patient enrichment strategy was implemented to increase the rate of SARS-CoV-2 positive participants enrolled at the CityHealth Urgent Care Oakland, CA site. For this strategy, patients presenting at the clinic were first tested by CityHealth Urgent Care staff, as part of the standard-of-care, with an FDA EUA rapid antigen test (Quidel Sofia SARS Antigen FIA) to pre-screen potential subjects prior to study enrollment. CityHealth Urgent Care staff were asked to identify patients with preliminary positive or negative results for study screening. The trained study operators, blinded to the original patient standard-of-care result, performed specimen collection, processing, and testing. An equal number of positives and negatives were pre-screened by the CityHealth Urgent Care staff at the CityHealth Urgent Care Oakland, CA site. Five unique study operators with varying healthcare backgrounds (licensed medical assistants and registered nurses) conducted the study between the two sites.

Between February 6, 2021 and March 9, 2021, study participants were enrolled at a third US clinical testing site (San Fernando Recreation Park in San Fernando, CA) who were at least 5 years of age and reported at least one of the COVID-19 symptoms listed above. Five unique study operators with varying healthcare backgrounds (licensed medical assistants and registered nurses) conducted the study at the San Fernando, CA site.

#### Testing procedure

Following the standard of care, patients were asked to provide a total of three nasal swab samples: a self-collected and observed nasal swab sample, followed by a second and third nasal swab sample that were collected by the health care provider (HCP). For the self-collected sample, the HCP provided specimen collection instructions and observed the specimen collection by the patient. The order of the second and third HCP-collected samples was randomized for testing with the investigational antigen test and a comparator method to ensure that bias was not introduced due to unequal distribution of viral material. Immediately after sample collection, the samples for the RT-PCR reference method were stored in viral transport media, while the other two nasal swabs were tested directly with the INDICAID™ Rapid Test according to the instructions for use (IFU).

INDICAID™ Rapid Test samples were tested immediately on-site after collection with no storage in accordance with the manufacturer’s protocol. Results of the INDICAID™ Rapid Test were interpreted by the test operators and recorded as “Positive,” “Negative”, or “Invalid” based on the visual presence or absence of the control and test lines on the developed test strip. Test results were recorded after 20 minutes of assay development by one individual operator per participant.

At the Oakland, CA and San Francisco, CA study sites, swab samples collected for RT-PCR were transferred to DNA/RNA Shield (Zymo Research, Irvine, CA), stored at room temperature, and shipped overnight at ambient temperature to Curative, Inc. (San Dimas, CA) per the manufacturer’s protocol for laboratory testing. The Curative SARS-CoV-2 Assay, an FDA Emergency Use Approved (EUA) test, was used as the RT-PCR reference method [10].

At the San Fernando, CA study site, swab samples collected for RT-PCR were transferred to BioCollections VTM (BioCollections Worldwide, Inc., Miami, FL), stored on ice packs, and shipped overnight on ice packs to BioCollections Worldwide, Inc. per the manufacturer’s protocol for laboratory testing. BioCollections first performed the FDA EUA assay Hologic Aptima SARS-CoV-2 to determine the SARS-CoV-2 status of each patient sample. The remaining volume from positive specimens were then stored frozen at -70°C and later analyzed with the FDA EUA BioCollections Worldwide SARS-CoV-2 Assay to obtain cycle threshold (Ct) results.

#### Ethical approval

To protect the rights, safety, and welfare of subjects, the study was conducted in accordance with 21 CFR 50, Protection of Human Subjects. Prior to study enrollment, each subject was asked to voluntarily provide his or her oral consent after being provided the IRB-approved Research Information Sheet. The onsite study investigator explained the nature, purpose, expected duration, and risks of study participation. Each potential subject had the opportunity to ask questions and receive answers from personnel conducting the study. All subjects were provided a copy of the Research Information Sheet. The IRB was reviewed by Advarra, IRB number: Pro00047510.

#### Statistical analysis

Difference testing for comparisons of groups in the population characteristics was performed with Chi-square testing for categorical variables. Positive percentage agreement (PPA), negative percentage agreement (NPA), overall percentage agreement (OPA), and accuracy with 95% confidence intervals were calculated using Wilson-Score method. RT-PCR results were used as the reference test. Analyses were performed on StataSE (StataCorp, College Station, TX). Figures were produced on GraphPad Prism 9.0.0 (GraphPad Holdings, La Jolla, CA).

### COVID-19 outbreak screening with the INDICAID™ COVID-19 Rapid Antigen Test

In this Dual-Track testing approach, the INDICAID™ Rapid Test is used to identify preliminary positives to trigger prioritization of sample processing for subsequent RT-PCR confirmatory testing. The outbreak testing was a collaborative effort between PHASE Scientific, ONCO Medical Laboratory, Hong Kong Sanatorium & Hospital, and the Food and Health Bureau of Hong Kong.

#### Outbreak testing population and locations

Twelve emergency outbreak testing centers (from December 10, 2020 to February 1, 2021) were organized at select locations in Hong Kong (Table 1). The sites were made available for asymptomatic individuals who perceived themselves as having a higher risk of exposure to SARS-CoV-2 or who were under compulsory testing requirements according to the guidelines from the Department of Health of the Government of the Hong Kong Special Administration Region.

**Table 1.** Dual-Track Testing Locations in Hong Kong

| **Approach A Locations** | **Approach B Locations** |
| --- | --- |
| Richland Gardens Apartments, Kowloon Bay | Maple Street Playground, Sham Shui Po |
| Tung Tau Estate, Chuk Un | Pei Ho Street Sports Center, Sham Shui Po |
| Jat Min Chuen Market, Shatin | Pitt Street location 1, Yau Ma Tei |
| Tsing Wah Playground, Tsing Yi | Pitt Street location 2, Yau Ma Tei |
| Tai Wo Hau Sports Center, Tai Wo Hau |  |
| Li Chi Kok |  |
| Kai Ching Estate, Kowloon City |  |
| Princess Margaret Hospital, Kwai Chung |  |

#### Dual-Track Testing Algorithms

Two Dual-Track testing algorithms were implemented (Figure 1). In both approaches, patient information is first collected at the registration station. Two nasal swab specimens and one oropharyngeal swab specimen are then collected by a clinician. One nasal swab specimen is used to perform the INDICAID™ Rapid Test immediately onsite. The additional nasal swab and oropharyngeal swab specimens are combined and stored in a single collection device containing viral transport medium (VTM) for subsequent RT-PCR testing.

**Figure 1.**
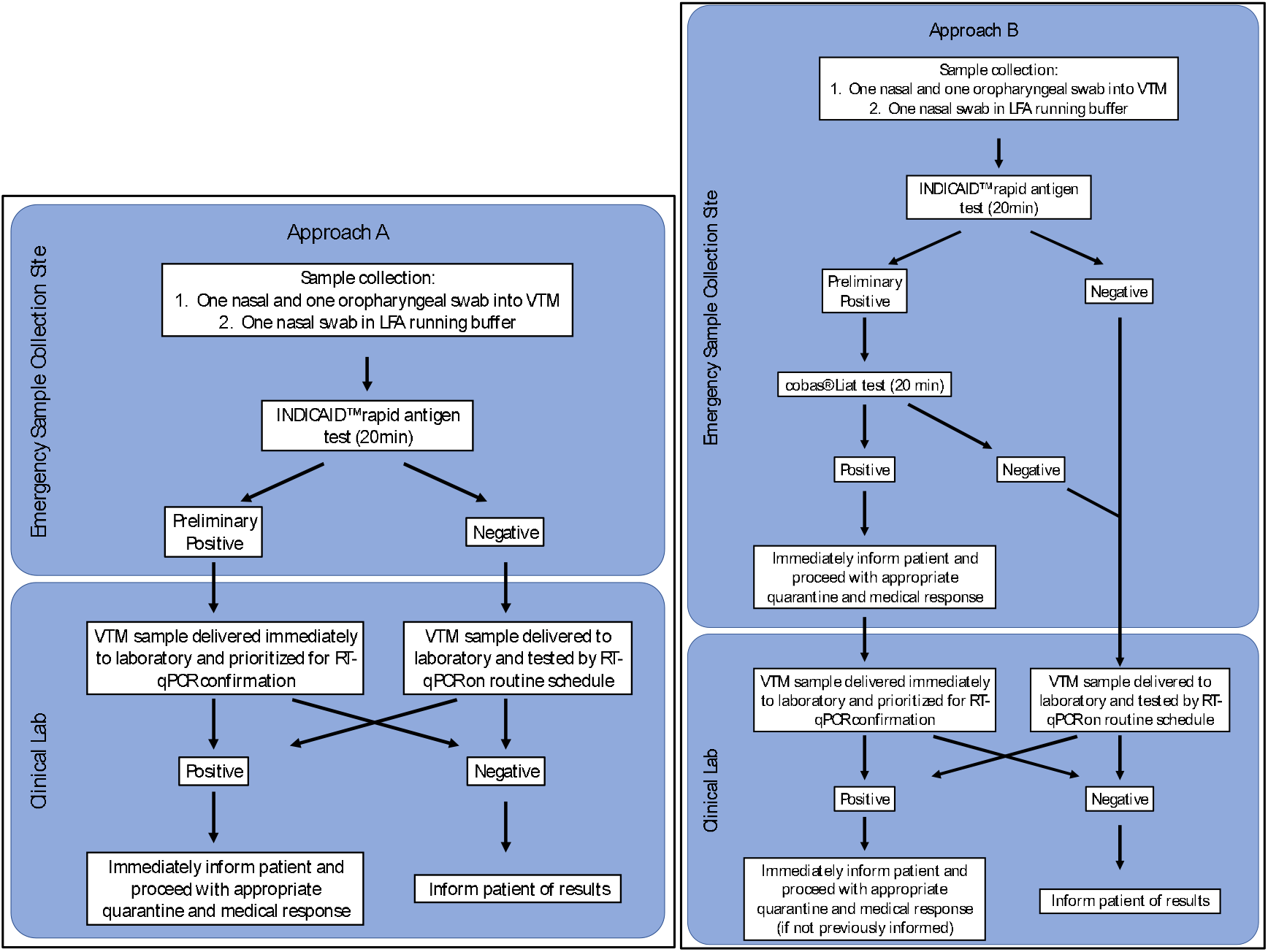
Flow charts of Dual-Track testing algorithm approaches A and B.

In Approach A, a preliminary positive result from the INDICAID™ Rapid Test would expedite the corresponding patient VTM sample for laboratory-based RT-PCR. Expedited testing would obtain results typically within an additional 8 hours compared to the standard approach requiring 48-hour government mandated turnaround time. In Approach B, a preliminary positive result from the INDICAID™ Rapid Test result would trigger the testing of the corresponding patient VTM sample with an onsite rapid nucleic acid amplification test (*cobas® SARS-CoV-2 & Influenza A/B Nucleic acid test on* the cobas® Liat system, Roche Molecular Diagnostics). Results from the onsite rapid nucleic acid amplification test would typically be obtained within an additional hour. In addition to the onsite rapid RT-PCR test, the corresponding patient VTM sample was also sent for expedited laboratory-based RT-PCR testing. All samples testing negative with the INDICAID™ Rapid Test were sent for RT-PCR testing with the standard approach by the ONCO Medical Laboratory in batches (Supplementary information).

For both approaches, a positive RT-PCR test resulted in the immediate notification of the patient to isolate and take precautionary measures according to the guidelines of the Department of Health of the Government of the Hong Kong Special Administration Region. The RT-PCR result turnaround time for positive samples detected using Approach A and B were evaluated at 8 and 4 different emergency sample collection sites located in Hong Kong, respectively (Table 1).

#### Statistical Analysis

Positive percentage agreement (PPA), negative percentage agreement (NPA), overall percentage agreement (OPA), and accuracy with 95% confidence intervals were calculated using Wilson-Score method. RT-PCR results were used as the reference test. Analyses were performed on StataSE (StataCorp, College Station, TX). Figures were produced on GraphPad Prism 9.0.0 (GraphPad Holdings, La Jolla, CA).

For both Dual-Track testing approaches and the standard testing approach, mean and standard deviation were calculated for the elapsed time (minutes) from sample collection to confirmatory RT-PCR test result. A two-tailed, two-sample T-test was performed comparing each of the Dual-Track approaches to the standard approach.

## RESULTS

### Prospective multi-site clinical evaluation of the INDICAID™ COVID-19 Rapid Antigen Test

#### San Francisco and Oakland population characteristics

In total, 83 participants with at least two COVID-19 symptoms were enrolled at the San Francisco, CA and Oakland, CA sites. Two participants were excluded from the analysis due to lost samples during transport for RT-PCR. Of the 81 participant specimens analyzed, 44.4% were from female participants (Table 2) with a median age of 32 years (IQR: 25, 44). The most frequently reported symptoms were muscle/body ache (61.7%), congestion/runny nose (60.5%), fatigue (56.8%), and headache (53.1%). The breakdown of duration of symptoms was 1-2 days in 37 participants (45.7%), 3-4 days in 38 participants (46.9%), and 5 days in 6 participants (7.4%).

**Table 2.**
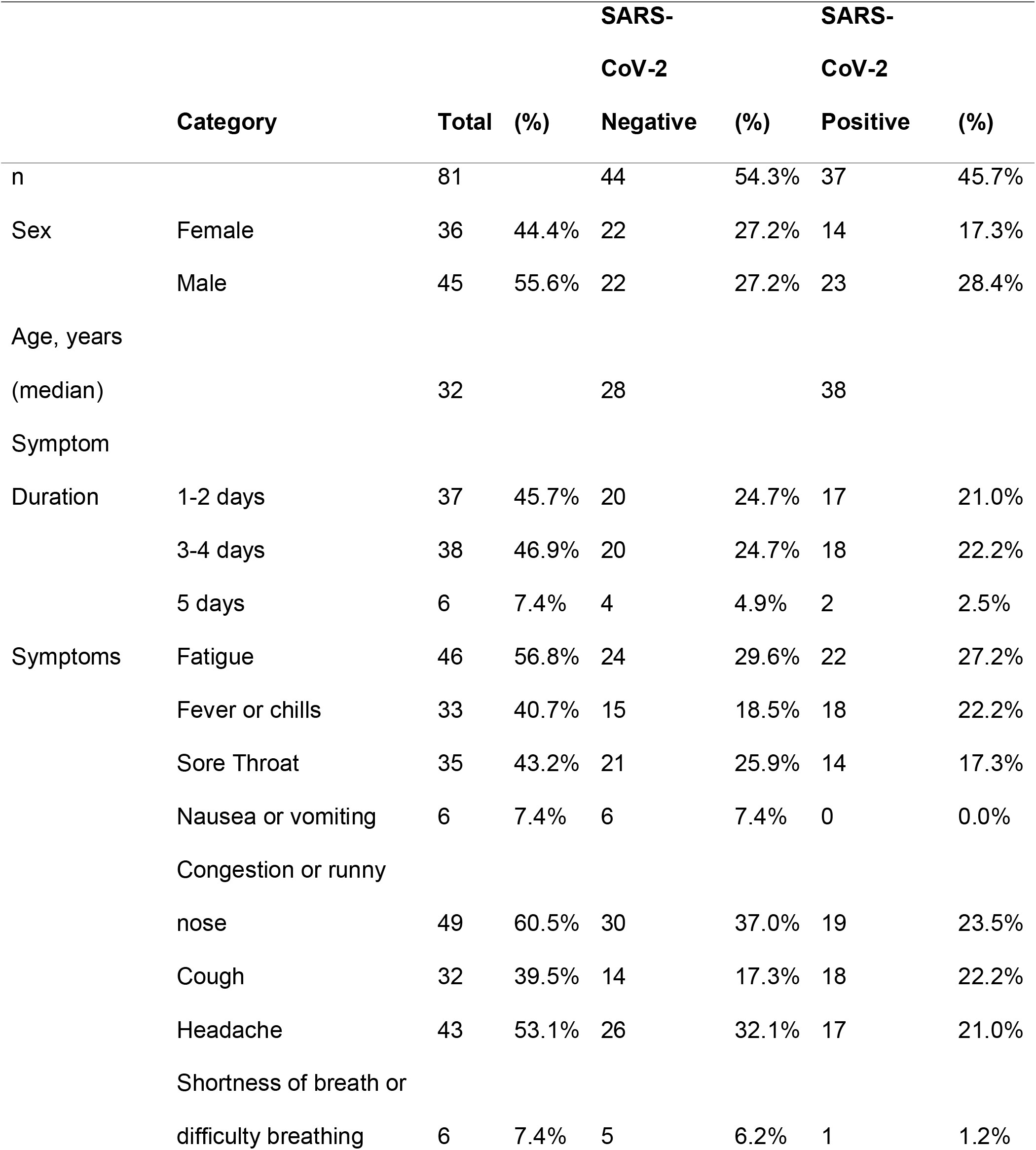
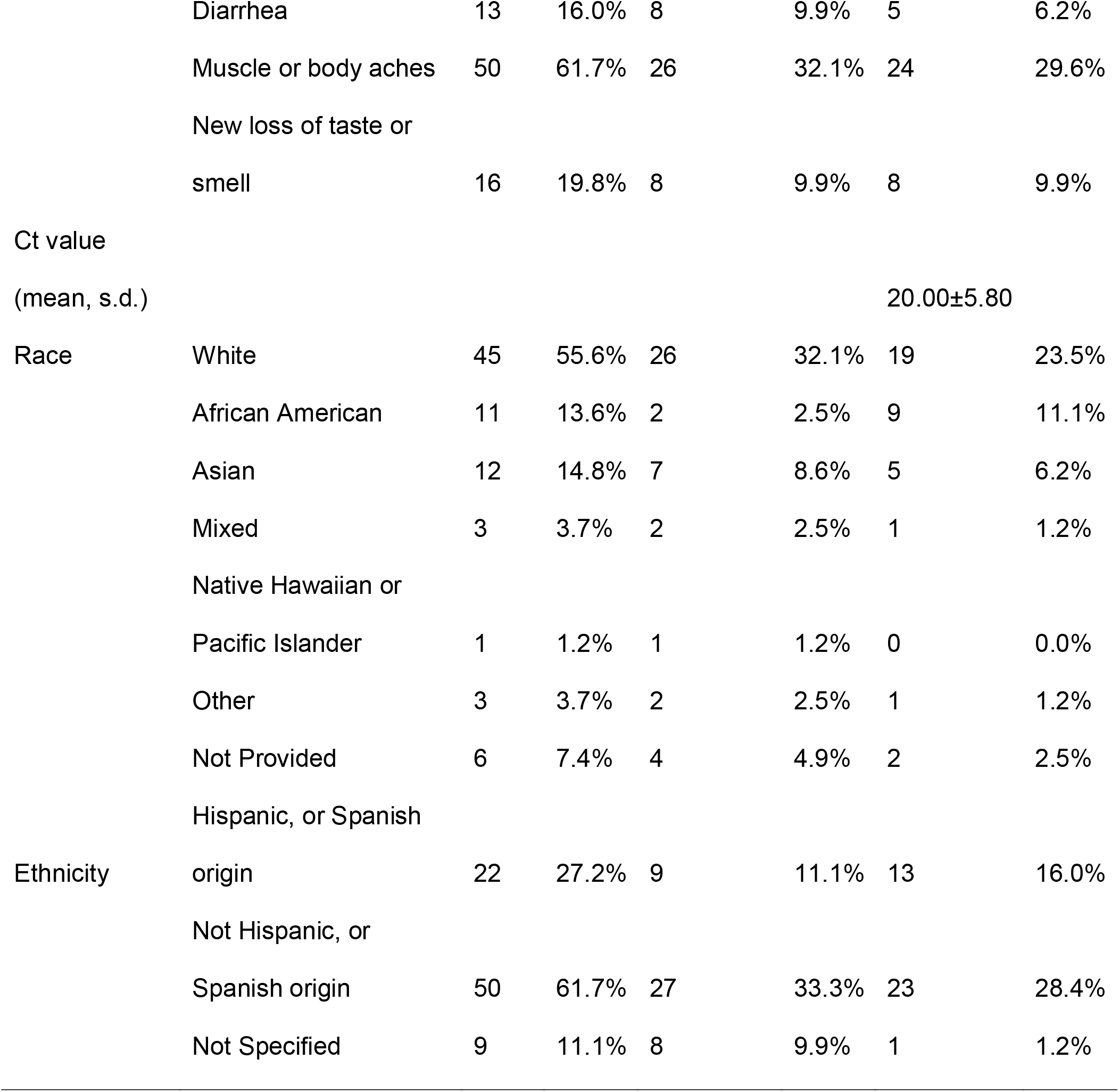
Population characteristics from San Francisco, CA and Oakland, CA study sites. Percentage values (%) represent the percentage out of the total 81 participants in the cohort.

#### San Fernando population characteristics

In total, 270 participants with at least one COVID-19 symptom were enrolled at the San Fernando, CA site. Two participants were excluded from the analysis due to lost or spilled samples during transport for RT-PCR. Of the 268 participant specimens analyzed, 52.6% were from female participants (Table 3) and the median age was 35 years (IQR: 24, 50). The most frequently reported symptoms were sore throat (60.8%), headache (60.1%), congestion/runny nose (59.0%), and cough (54.9%). The distribution of duration of symptoms was 1-2 days in 109 participants (40.7%), 3-4 days in 127 participants (47.4%), and 5 days in 32 participants (11.9%).

**Table 3.**
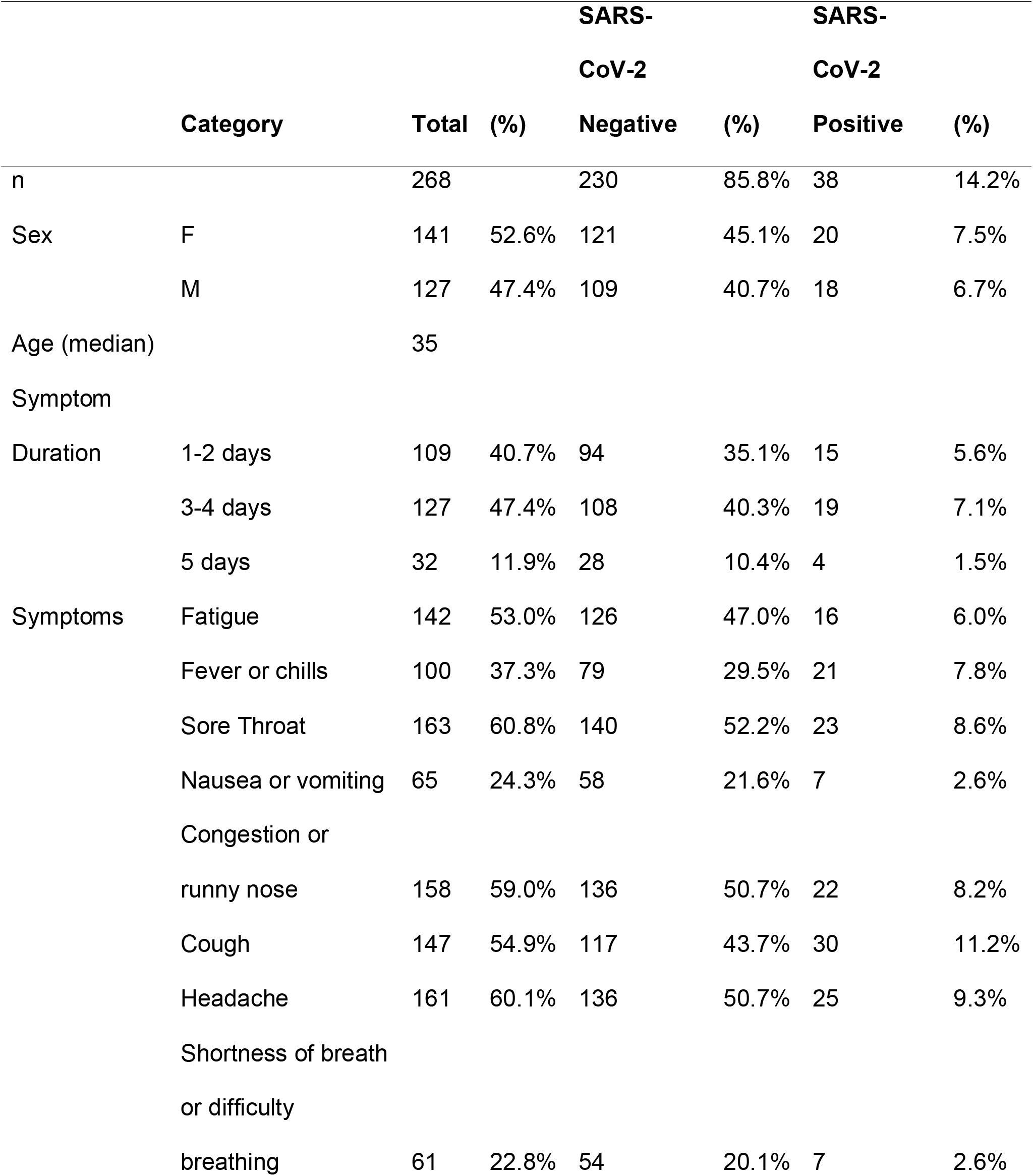
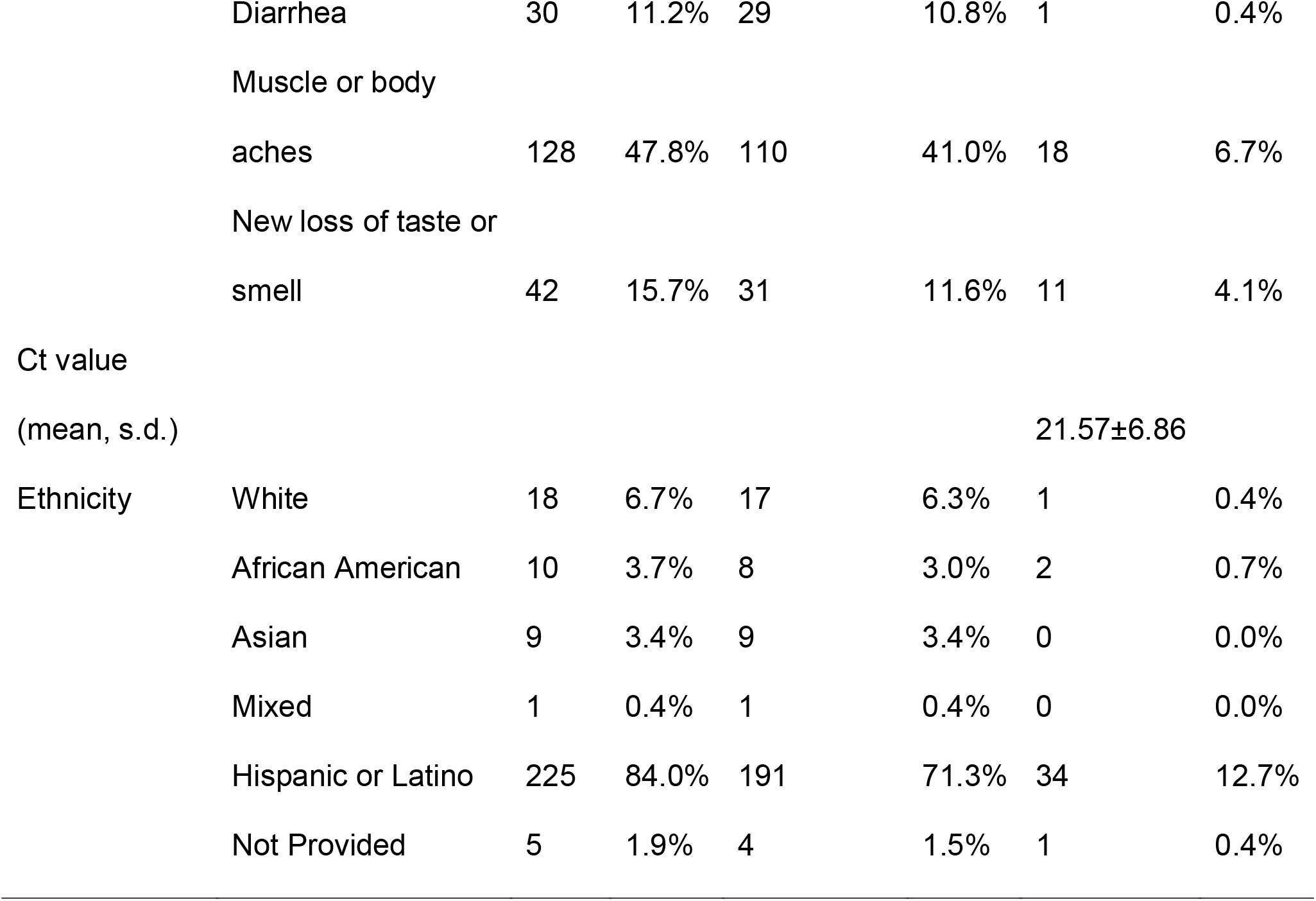
Population characteristics from the San Fernando, CA study site. Percentage values (%) represent the percentage out of the total 268 participants in the cohort.

#### Performance of the INDICAID™ COVID-19 Rapid Antigen Test in symptomatic patients

Of the total 329 participant specimens included in the analyses, 75 tested positive with the comparator laboratory-based RT-PCR test. The mean cycle threshold value was 20.79±6.39 (Figure 2). The INDICAID™ Rapid Test demonstrated a PPA of 85.3% (95% Confidence Interval [95% CI]: 75.6% - 91.6%) and NPA of 94.9% (95% CI: 91.6% - 96.9%) when sample collection was conducted by a healthcare professional.

**Figure 2.**
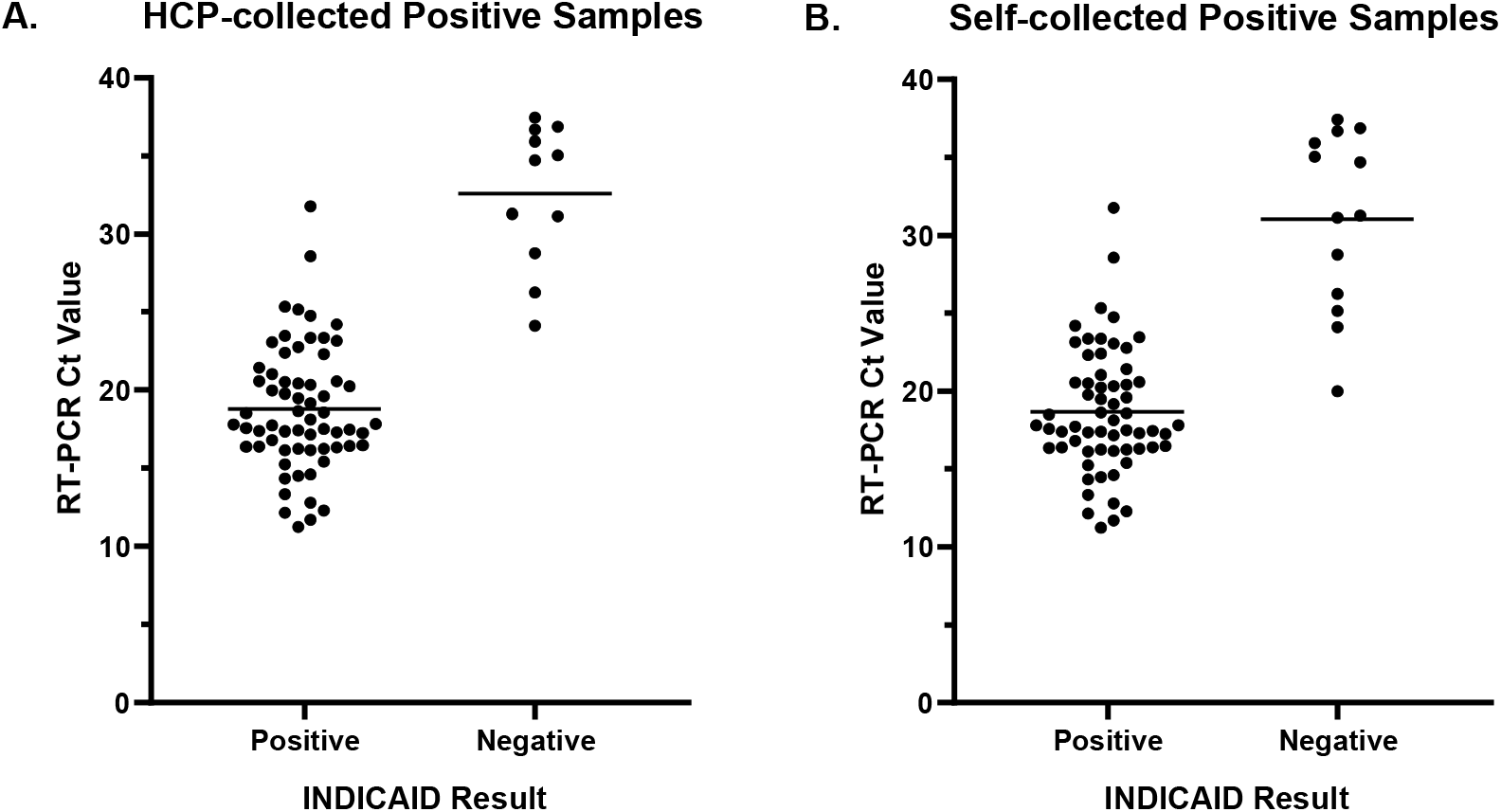
Association between Ct value and INDICAID™ Rapid Test result when using (A) healthcare professional-collected samples and (B) self-collected samples for INDICAID™ Rapid Test.

**Figure 3.**
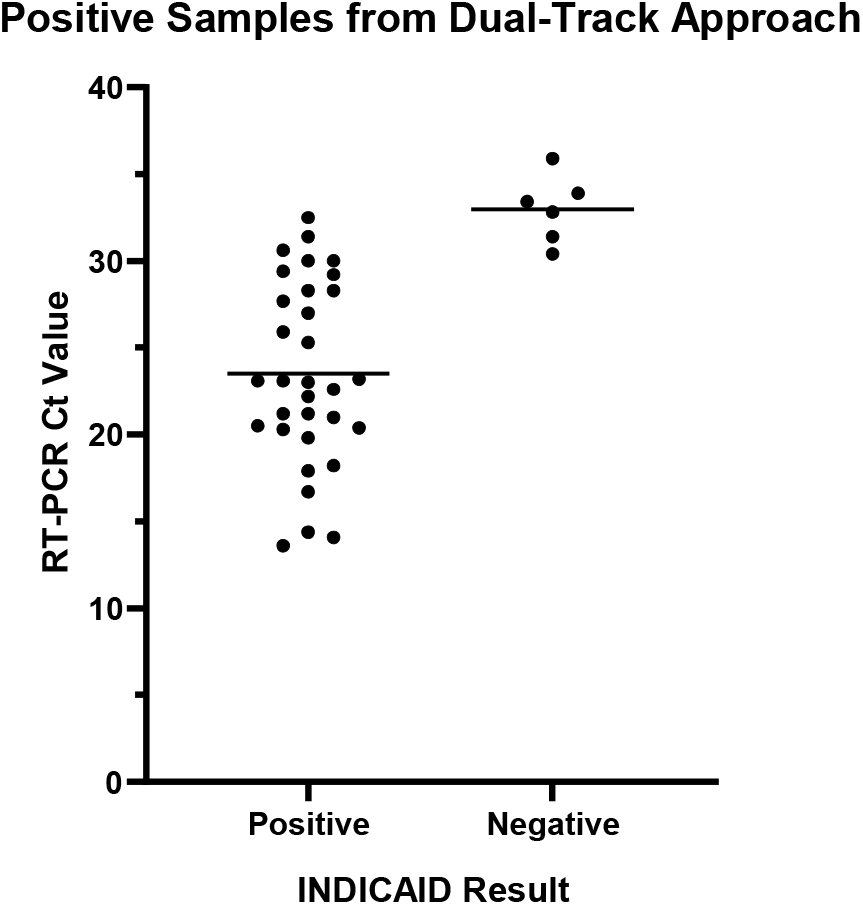
Association between Ct value and INDICAID™ Rapid Test results when implemented in outbreak screening centers.

When sample collection was conducted by the participant (self-collected), the PPA was 82.7% (95% CI: 72.6% - 89.6%) and the NPA was 96.4% (95% CI: 93.4% - 98.0%). OPA for HCP- and self-collected specimens were 92.8% and 93.4%, respectively (Tables 4 and 5).

**Table 4.** INDICAID™ COVID-19 Rapid Antigen Test Performance within 5 Days of Symptom Onset Against RT-PCR Reference Method (HCP-Collected Sample)

| **INDICAID™ COVID-19 Rapid**<br>**Antigen Test** | **Comparative Method (RT-PCR)** |  |  |
| --- | --- | --- | --- |
|  | **Positive** | **Negative** | **Total** |
| Positive | 64 | 14 | 78 |
| Negative | 11 | 260 | 271 |
| **Total** | 75 | 274 | 349 |

|  | **95% confidence interval** |  |  |
| --- | --- | --- | --- |
|  | **Percent** | **Low Limit** | **High Limit** |
| **Positive Percent Agreement (PPA)** | 85.3% | 75.6% | 91.6% |
| **Negative Percent Agreement (NPA)** | 94.9% | 91.6% | 96.9% |
| **Overall Percent Agreement (OPA)** | 92.8% | 89.6% | 95.1% |

**Table 5.** INDICAID™ COVID-19 Rapid Antigen Test Performance within 5 Days of Symptom Onset Against RT-PCR Reference Method (Self-Collected Sample)

| **INDICAID™ COVID-19 Rapid**<br>**Antigen Test** | **Comparative Method (RT-PCR)** |  |  |
| --- | --- | --- | --- |
|  | **Positive** | **Negative** | **Total** |
| Positive | 62 | 10 | 72 |
| Negative | 13 | 264 | 277 |
| **Total** | 75 | 274 | 349 |

|  | **95% confidence interval** |  |  |
| --- | --- | --- | --- |
|  | **Percent** | **Low Limit** | **High Limit** |
| **Positive Percent Agreement (PPA)** | 82.7% | 72.6% | 89.6% |
| **Negative Percent Agreement (NPA)** | 96.4% | 93.4% | 98.0% |
| **Overall Percent Agreement (OPA)** | 93.4% | 90.3% | 95.6% |

### COVID-19 outbreak screening with the INDICAID™ COVID-19 Rapid Antigen Test

#### Performance of the INDICAID™ COVID-19 Rapid Antigen Test in asymptomatic patients

In total, 22,994 asymptomatic individuals were screened at 12 outbreak screening centers in Hong Kong. Thirty-eight (38) of the total 22,994 patients tested positive for SARS-CoV-2 by laboratory-based RT-PCR. The INDICAID™ Rapid Test demonstrated a PPA of 84.2% and NPA of 99.9% against the comparator RT-PCR method (Table 6). All samples tested on-site by the *cobas® SARS-CoV-2 & Influenza A/B Nucleic acid test* demonstrated concordant results with laboratory-based RT-PCR from ONCO Medical Laboratory.

**Table 6.** INDICAID™ COVID-19 Rapid Antigen Test Performance During Implementation in Outbreak Screening Centers in Hong Kong

| **INDICAID™ COVID-19 Rapid Antigen Test** | **Comparative Method (RT-PCR)** |  |  |
| --- | --- | --- | --- |
|  | **Positive** | **Negative** | **Total** |
| Positive | 32 | 18 | 50 |
| Negative | 6 | 22,938 | 22,944 |
| **Total** | 38 | 22,956 | 22,994 |
|  | **95% confidence interval** |  |  |
|  | **Percent** | **Low Limit** | **High Limit** |
| **Positive Percent Agreement (PPA)** | 84.2% | 69.6% | 92.6% |
| **Negative Percent Agreement (NPA)** | 99.9% | 99.9% | 100.0% |
| **Overall Percent Agreement (OPA)** | 99.9% | 99.8% | 99.9% |

#### Time to positive result confirmation for each Dual-Track testing algorithm

Time-to-result data from 8 of 10 positive samples in Approach A and 12 of 18 positive samples in Approach B were available for analyses. The time-to-result for the standard approach was estimated using 299 negative samples randomly selected across three days and multiple sites as the extraction of reported time data from all 22,944 negative samples would be a labor-intensive process.

After a preliminary positive result with the INDICAID™ Rapid Test, both Approach A and Approach B demonstrated a shorter time to positive confirmatory RT-PCR result when compared to the standard approach. Positive patient confirmation for Approach A was on average 7.0 hours (mean=420±151 min) compared to an average of 10.85 hours (mean=651±171 min) for the standard approach (t(7)=4.3, p<0.004). Similarly, for Approach B, positive patient confirmation was on average 0.84 hours (mean=50.4±14 min) compared to an average of 10.85 hours (mean=651±171 min) for the standard approach (t(232)=56.3, p<0.0001).

## DISCUSSION

In the prospective clinical study in symptomatic patient populations, the INDICAID™ Rapid Test demonstrated a PPA of 85.3% and NPA of 94.9% against comparator laboratory-based RT-PCR. False negative results were mainly observed in participant specimens with higher Ct values and likely lower viral loads. Although participant collected (self-collected) samples resulted in slightly lower performance compared to clinician collected samples, self-collection allows for significantly reduced risk to healthcare workers and improved clinical workflow for high throughput screening operations.

When implemented for outbreak testing of asymptomatic patients, the INDICAID™ Rapid Test demonstrated a PPA and NPA of 84.2% and 99.9%, respectively, against laboratory-based RT-PCR. The similar PPA and NPA compared to the prospective clinical study suggest minimal or no loss in performance when used in asymptomatic populations in a real-world setting. When combined with RT-PCR testing in the Dual-Track testing approach, the INDICAID™ Rapid Test was able to successfully reduce the time to confirmatory positive result from an average of 10.85 hours to 0.84 hours. The reduced time to positive patient identification and notification could result in a significant reduction in transmission in densely populated communities.

There were limitations during the symptomatic clinical evaluation. The tests were conducted under controlled temperature and lighting conditions and interpreted by a limited pool of trained operators. Widespread community testing might occur under less-controlled conditions. Several point-of-care flex studies (see Supplementary Information) were performed to demonstrate that the INDICAID™ Rapid Test is robust under suboptimal conditions (i.e., extreme temperature and humidity, out-of-specification buffer addition, and variable result reading times). Additionally, self-collected samples were always collected first, which might have affected the amount of virus/antigen remaining for the subsequent operator-collected samples. While the sample collection order is not expected to drastically influence the reference test result, the effect of repeated sampling on the INDICAID™ Rapid Test has not been confirmed.

Rapid antigen tests are expected to play an increasingly important role in COVID-19 testing programs. To our knowledge, our Dual-Track testing approach is one of the first widely coordinated efforts to integrate COVID-19 rapid antigen testing with rapid confirmatory RT-PCR testing in asymptomatic populations. Recently, the United States CDC proposed similar screening algorithms that incorporate routine rapid antigen testing for asymptomatic populations, followed by positive patient confirmation by RT-PCR [11]. The performance of the INDICAID™ COVID-19 Rapid Antigen Test makes it a suitable test to be incorporated into such screening algorithms as communities, schools and businesses reopen with the relaxation of public health measures. The Dual-Track testing approach may be an effective solution for COVID-19 surveillance and outbreak response, particularly in densely populated communities.

## CONCLUSION

The INDICAID™ COVID-19 Rapid Antigen Test demonstrated high PPA and NPA against laboratory-based RT-PCR in symptomatic and asymptomatic populations with varying community rates of circulating SARS-CoV-2. Furthermore, when used in an algorithm testing approach (Dual-Track), the INDICAID™ COVID-19 Rapid Antigen Test successfully reduced the time to confirmatory positive result. The Dual-Track testing approach highlights the advantages that rapid antigen testing can bring to already established RT-PCR testing frameworks. More studies are needed to determine optimal use of rapid antigen testing for screening algorithms.

## Data Availability

Data is available by request if requests are reasonable.

## ACKNOWLEDGMENTS

The authors would like to thank the Hong Kong SAR Government and the Hong Kong Food and Health Bureau for the opportunity to integrate the Dual-Track testing approach into the Hong Kong emergency COVID-19 testing program and for their support in the evaluation and feedback on the performance of the Dual-Track testing approach. We thank The University of Hong Kong for their support in the evaluation of the Dual-Track testing performance data. The authors would also like to thank the study staff at CityHealth Urgent Care, City of San Fernando and the San Fernando Fire Department who contributed to the success of this study.

## Supplementary Information

### ONCO Medical Laboratory RT-PCR Clinical laboratory testing procedure

Total viral RNA was extracted from 200µl of patient VTM sample using either pre-filled 96-deep well plate (64T, Tianlong Technology) with automated GeneRotex 96 rotary nucleic acid extractor system (NANBEI, Tianlong Technology), or 96-well pre-packed extraction reagents (SDK60104-96T, Bioperfectus Technologies) with automated nucleic acid extraction system (SSNP-3000A, Bioperfectus Technologies). RT-PCR was performed to determine the expression level of *Orf1b* in the extracted RNA using PHASIFY™ DeCOVID SARS-CoV-2 RT-qPCR Kit (3010100, Phase Scientific) according to the manufacturer’s protocol. Sample quality was validated via measuring expression levels of internal controls (viral: *RdRP*; human: *RNase P*). Positive and negative controls were included in each PCR reaction.

### Analytical Validation of the INDICAID™ COVID-19 Rapid Antigen Test Materials and Methods

#### Analytical limit of detection

The limit of detection (LoD) was determined by limiting dilution studies using characterized gamma-irradiated SARS-CoV-2 virus (BEI Resources, NIAID, NIH, SARS-Related Coronavirus 2, Isolate USA-WA1/2020, Gamma-Irradiated, NR-52287) spiked into pooled human nasal matrix from healthy donors (IRHUNF1ML, Innovative Research, MI, USA). At each dilution, 50 μL of sample was inoculated onto swabs and then assayed using the INDICAID™ COVID-19 Rapid Antigen Test procedure. An initial range finding study was performed using a 10-fold dilution series of the characterized SARS-CoV-2, testing the device in triplicate at each concentration. Concentrations between the last dilution that produced three positive test results and the first dilution to produce at least one negative test result were further evaluated using a 2-fold dilution series, in triplicate for each level, to refine the tentative LoD. This LoD was then confirmed by testing 20 replicates with concentrations at the refined tentative limit of detection. The final LoD of the test was determined to be the lowest concentration resulting in positive detection of at least 19 out of 20 replicates.

To correlate the performance of the INDICAID™ Rapid Test with cycle threshold value output, 14 concentrations of SARS-CoV-2 from heat inactivated SARS-Related Coronavirus 2 Culture Fluid (1.02x 10^8^ TDID_50_/mL, 0810587CFHI, lot 325309, Zeptometrix) were prepared through serial dilution in the INDICAID™ Rapid Test buffer and then tested with both the INDICAID™ Rapid Test and with *ONCO Medical Laboratory RT-PCR method*. A dilution scheme that simulated the differences in dilution ratios between the INDICAID™ Rapid Test and the ONCO Medical Laboratory RT-PCR test was used. Viral-free test buffer was included as negative controls. The INDICAID™ Rapid Test was performed using 75µl of sample immediately after the dilution. Positive and negative band determinations were made by visual inspection from three blinded observers according to a standardized line intensity reference chart. Tests were analyzed at 20 minutes.

To determine the RT-PCR cycle threshold vales of the contrived samples, total viral RNA was extracted from 200µl of sample using 96-well pre-packed extraction reagents (SDK60104-96T, Bioperfectus Technologies) with automated nucleic acid extraction system (SSNP-3000A, Bioperfectus Technologies). The expression level of *Orf1b* in the extracted RNA was determined using PHASIFY™ DeCOVID SARS-CoV-2 RT-qPCR Kit (3010100, Phase Scientific) according to the manufacturer’s protocol. Sample quality was validated via measuring expression levels of internal controls (viral: *RdRP*; human: *RNase P*). Positive and negative controls were included in each PCR reaction. Non-linear regression analysis was performed on GraphPad Prism 9.0.0 fit to a sigmoidal curve constraining the top plateau at a rapid antigen test line intensity 12.

#### Endogenous interference, cross-reactivity, microbial interference

After the LoD was determined, evaluations of endogenous interference, cross-reactivity, and microbial interference were conducted according to the US FDA’s Emergency Use Authorization (EUA) template for SARS-CoV-2 antigen test manufacturers.[3]

#### Flex studies for out-of-specifications test performance

A thorough hazard analysis was conducted to evaluate the impact of errors, or out-of-specifications conditions, on the rapid antigen test performance. To test the effect of extreme environmental conditions, contrived samples of 5.6 × 10^3^ TCID_50_/mL (2x the determined analytical LoD) gamma-irradiated SARS-CoV-2 in pooled nasal matrix, as well as non-spiked negative pooled nasal matrix, were tested on the INDICAID™ Rapid Test in low temperature (2-8°C) and high temperature/high humidity (40°C and near 95% relative humidity (RH)) conditions. One hour prior to the study, test kits were placed in a refrigerator maintaining 2-8°C or an incubator maintaining 40°C and 95% RH. Contrived samples were then applied to the test devices and allowed to run for 20 min in the same respective environments (n=3 per condition). Test results were recorded after 20 min.

To test the effect of INDICAID™ Rapid Test buffer volume variability, contrived samples of 5.6 × 10^3^ TCID_50_/mL gamma-irradiated SARS-CoV-2 in pooled nasal matrix, as well as non-spiked negative pooled nasal matrix, were tested on the INDICAID™ Rapid Test at room temperature. Following the release of contrived specimen from the inoculated swab into the buffer solution, solution volumes of 1, 2, 3, 4, 5, 6 drops, and the entire buffer volume were applied to the test device (n=3 per condition). Test results for all replicates were interpreted at 20 minutes.

To test the effect of variable result read times, contrived samples of 5.6 × 10^3^ TCID_50_/mL gamma-irradiated SARS-CoV-2 in pooled nasal matrix, as well as non-spiked negative pooled nasal matrix, were tested on the INDICAID™ Rapid Test at room temperature. Test results were interpreted at 5, 10, 15, 20, 30, and 60 minutes after samples had been applied to the test device (n=3 per condition).

## RESULTS

### Analytical limit of detection

For the initial LoD range finding study, 10-fold serial dilutions of gamma-irradiated SARS-CoV-2 in pooled human nasal matrix were prepared with the highest test concentration of 2.8 x10^5^ TCID_50_/mL (1.4 × 10^4^ TCID_50_/swab). From this dilution series, the lowest concentration to produce 3 out of 3 positive results on the INDICAID™ Rapid Test was 2.8 x10^3^ TCID_50_/mL (1.4 × 10^2^ TCID_50_/swab). This tentative LoD was further refined using 2-fold serial dilutions between 2.8 x10^3^TCID_50_/mL (1.4 × 10^2^ TCID_50_/swab) and 1.75 × 10^2^ TCID_50_/mL (8.75 TCID_50_/swab). From this 2-fold dilution series, a concentration of 2.8 × 10^3^ TCID_50_/mL (1.4 × 10^2^ TCID_50_/swab) continued to be the lowest concentration that produced 3 out of 3 positive results. This concentration was confirmed to be the final LoD as 20 out of 20 replicates produced a positive result with test samples containing 1.4 × 10^2^ TCID_50_/swab.

The LoD in relation to cycle threshold value of the INDICAID™ Rapid Test was evaluated using contrived samples of varying concentrations of heat inactivated SARS-CoV-2 spiked into the INDICAID™ Rapid Test buffer. Results were reported in line intensity values from a standardized line intensity reference chart by three trained readers in a blind experimental design. Line intensity values correspond to the visibility of the test line to the user with 0 representing no visible test line and 12 representing the maximum test line intensity (Figure S1). Based on an industry standard for visually based lateral-flow immunoassays, a line intensity of 3 was utilized as the visual cut-off for the intended user (non-laboratory healthcare professionals). The visual cut-off intersects the non-linear regression line (R^2^ = 0.976) at a Ct value of 27.2.

**Supplementary Figure S1.**
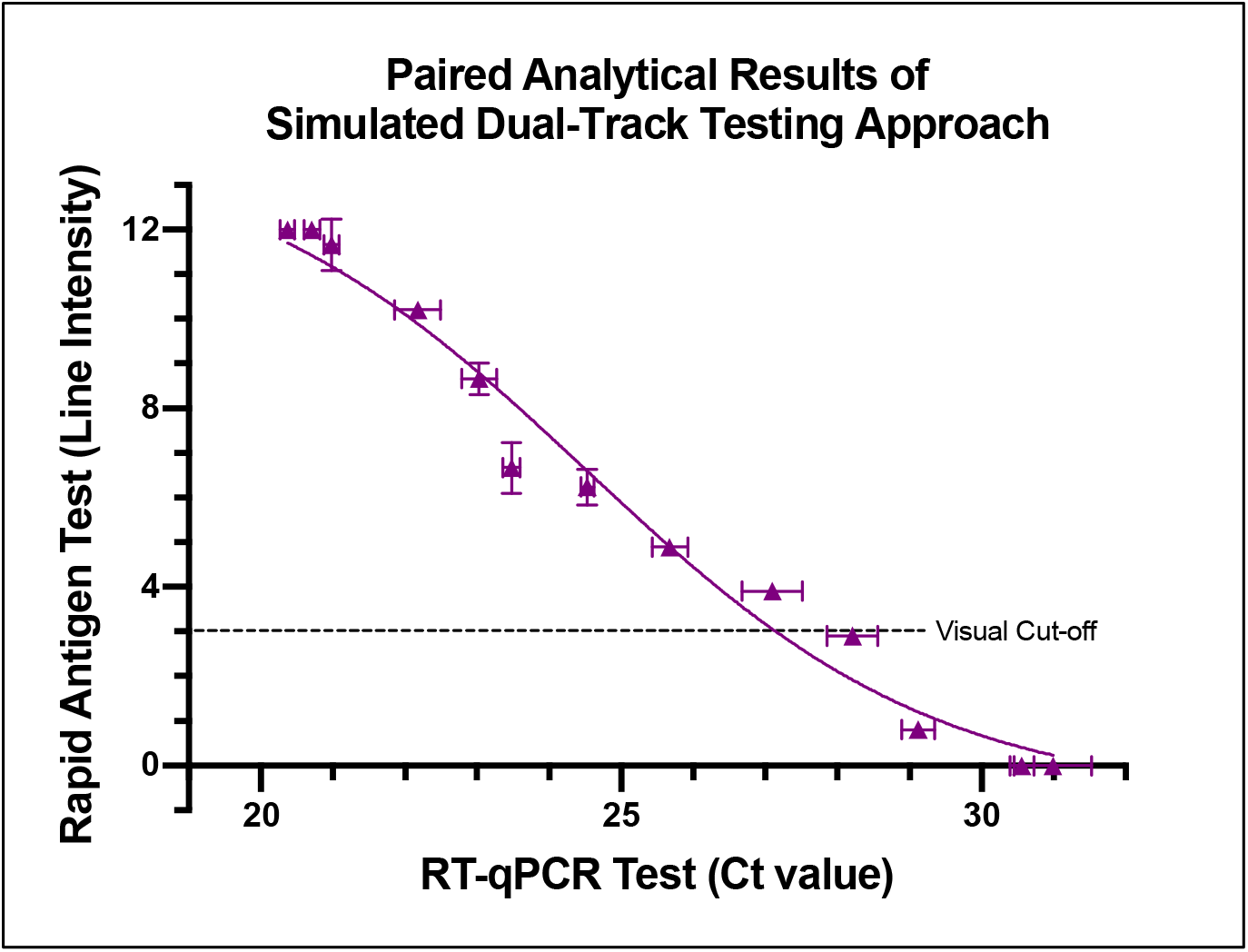
Correlation of INDICAID™ COVID-19 Rapid Antigen Test and RT-PCR Ct results. Non-linear regression analysis was performed (R^2^ = 0.976). All error bars are standard deviation.

#### Endogenous interference, cross-reactivity, microbial interference

No cross-reactivity nor test interference were observed for 27 common respiratory pathogens and pooled nasal wash in the presence of gamma-irradiated SARS-CoV-2 at 3x the analytical LoD. Furthermore, 14 endogenous substances that may be found in respiratory specimens of patients symptomatic for respiratory illness demonstrated no significant test interference in the presence of gamma-irradiated SARS-CoV-2 at 3x the analytical LoD.

#### Flex studies for out-of-specifications test performance

A series of flex studies was conducted to evaluate the influence of errors that can occur in point-of-care environments. These errors include extreme temperatures, extreme humidity, higher- and lower-than-recommended buffer volumes added to the test device, and sooner- and later-than-recommended read times. When tested with contrived samples in low temperature conditions (2-8°C) as well as high temperature and humidity conditions (40°C and 95% RH), the INDICAID™ Rapid Test produced expected positive and negative results and no invalid test results were observed.

With low positive contrived samples (i.e., 2x the analytical LoD), accurate test result interpretation could be made by trained users as soon as 10 min and as late as 60 min after samples have been applied to the test device. Furthermore, accurate results were produced when 2-6 drops of the INDICAID™ Rapid Test buffer are applied to the test device, as opposed to the manufacturer-recommended 3 drops.

